# Caesarean delivery patterns and selected maternal-neonatal outcomes in an urban Tanzanian referral hospital: a retrospective record review

**DOI:** 10.64898/2026.11.16.26368240

**Authors:** Grace Watts, Vincent Chambo, Luzango Maembe, Taibar Haidar, Zavery Benela, Revocatus Kabanga

## Abstract

**Background:** Caesarean delivery can be lifesaving when clinically indicated, but high facility rates require regular audit to distinguish appropriate emergency obstetric care from potentially avoidable operative delivery. This study described caesarean delivery patterns and selected maternal-neonatal outcomes among women delivering at Mwananyamala Regional Referral Hospital in Dar es Salaam, Tanzania.

**Methods:** A retrospective record review was conducted using complete delivery records from January 2025 to June 2025. The analytic dataset included 302 eligible records abstracted from delivery registers, admission records, and neonatal logs. The primary outcome was caesarean delivery. Associations were assessed using logistic regression restricted to variables available before or at the time of delivery decision. Post-delivery variables were not treated as predictors of caesarean delivery. Gestational-age values below 28 weeks or missing values were treated as invalid/missing and excluded from the study.

**Results:** Mean maternal age was 28.09 ± 6.68 years; 9.93% were <20 years, 68.87% were 20–34 years and 21.19% were ≥35 years. Most women were multiparous (53.3%) and 43.71% delivered by caesarean section. In multivariable analysis, caesarean delivery was associated with employment (AOR=3.54; P<0.001), primigravity (AOR=2.65; P=0.009), prior uterine scar (AOR=1.89; P=0.002), and high-risk pregnancies (AOR=2.68; P=0.003). Maternal complications were infrequent, but postpartum hemorrhage was more common among older mothers. Neonatal outcomes were largely favorable, with 92.4% live births and 15.2% low birth weight.

**Conclusion:** Caesarean delivery was common among complete records at this urban referral hospital, but maternal age was not an independent predictor. The results support routine caesarean- delivery audit, improved delivery-register documentation, and future Robson-classification analysis once the necessary variables are captured reliably.

**What is already known –**Maternal age, parity, high-risk pregnancy, previous caesarean delivery, and gestational age influence obstetric and neonatal outcomes. Facility-level caesarean delivery rates are increasingly used as markers of obstetric service utilization and quality, but crude rates are difficult to interpret without structured audit and adequate routine-data quality.

**What this study adds** – This study found a high caesarean delivery rate (43.7%) and identified primigravity, prior uterine scar, and maternal comorbidities as predictors of caesarean delivery. Older maternal age was associated with increased postpartum hemorrhage, while stillbirth remained an important neonatal concern. The findings provide recent hospital-based evidence from an urban Tanzanian setting.

**How this study might affect research, practice or policy** – The findings highlight the need for improved intrapartum monitoring, targeted management of high-risk pregnancies, and regular audits of caesarean delivery indications. Mwananyamala Regional Referral Hospital should strengthen delivery-register completeness, capture valid previous caesarean history for all women, and add missing variables needed for Robson Ten-Group Classification, especially fetal presentation or lie and number of fetus(es). The study also provides baseline data for future maternal health research.

## Introduction

Maternal and neonatal outcomes remain public health priorities. Despite global progress, approximately 260,000 women died in 2023 from pregnancy or childbirth-related causes[1], and about 2.3 million newborns died during their first month of life[2]. In Sub-Saharan Africa, the adolescent (aged 15–19) birth rate exceeds 90 per 1,000 [3]. Such pregnancies carry higher risks of obstetric complications and poor neonatal outcomes[4]. At the other extreme, advanced maternal age (AMA, ≥35 years) is rising globally: WHO surveys estimate ∼12.3% of pregnancies in developing countries are AMA[5]. AMA has been consistently linked to adverse outcomes: caesarean delivery, gestational diabetes, hypertension, and fetal growth restriction[6,7].

Gravidity and parity have also been known to shape obstetric risk. Primigravity is a known risk factor for hypertensive disorders and labor dystocia. Conversely, grand multiparity has been associated with placenta-related complications, postpartum hemorrhage, gestational diabetes, and perinatal mortality[8–10]. However, crude age and parity associations alone are rarely sufficient to guide facility-level quality improvement. In referral hospitals, caesarean delivery rates may be influenced by case mix, previous caesarean delivery, high-risk pregnancy, gestational age, fetal presentation, labour onset, referral status, provider practice, and data quality.

The initial framing of this manuscript as a broad study of maternal age, parity, and obstetric outcomes was reconsidered because the topic was well established and the earlier analysis included post-delivery variables as predictors of caesarean delivery. Therefore, the study was reframed to reflect caesarean delivery patterns and selected maternal-neonatal outcomes in an urban Tanzanian referral hospital.

## Main Objective

- To describe caesarean delivery patterns among women delivering at Mwananyamala Regional Referral Hospital.

## Specific Objectives

- To determine the incidence of adverse obstetric outcomes across different maternal age groups among women delivering at Mwananyamala Regional Referral Hospital.
- To describe the distribution of neonatal outcomes among women delivering at Mwananyamala Regional Referral Hospital
- To investigate the factors associated with caesarean delivery using a corrected analysis restricted to variables available before or at the time of delivery decision.

## Methods

### Study design and setting

A retrospective cross-sectional study was conducted at Mwananyamala Regional Referral Hospital, an urban referral hospital in Dar es Salaam. Data were accessed and collected from 15^th^ July 2025 to 8^th^ August 2025, from delivery registers, admission records, and neonatal logs (S1_Data) corresponding to deliveries that occurred from January to June 2025. A consecutive sampling technique was used, whereby all eligible delivery records available during the study period were reviewed and screened for inclusion in the study. Records were extracted from both vaginal delivery and caesarean delivery registers covering January to June 2025. Following data cleaning and verification, only records meeting the inclusion criteria and containing complete information on key study variables were included in the final analysis.

### Eligibility criteria

All women who delivered at Mwananyamala Regional Referral Hospital (MRRH) during the study period were eligible for inclusion in the study. Only records with documented maternal age at admission, parity, mode of delivery, intrapartum complications, and APGAR score were included in the analysis. Women with multiple gestations, major fetal anomalies, or missing information on key study variables (such as maternal age, parity, or mode of delivery) were excluded from the study(Fig_1).

## Study variables

Variables were conceptualized using a framework adopted from a study conducted in Rwanda [13] and a data collection tool was developed and tested using a pilot data set before its introduction into the field.

A Kaiser-Meyer-Olkin (KMO) test was used to evaluate sampling adequacy as well as the suitability of conducting a factor analysis in our extracted data. After confirming factorability, a principal axis factoring (PAF) extraction method was employed and the overall KMO value was 0.7 (indicating sampling adequacy) (S1_Text).

Dependent variables were:

Mode of delivery (whether by Caesarean Delivery (CD) or Spontaneous Vertex Delivery (SVD)) as documented i.e., SVD (unassisted or assisted), Elective CD (planned prior to labor onset) and Emergency CD (performed after labor onset for maternal or fetal indication).

Independent variables were categorized into:

Socio-demographic characteristics: Maternal age groups (teenagers: < 20 years, reference: 20–34 years and advanced age: ≥ 35 years), place of residence, marital status, education level, occupation, health insurance and referral status Obstetric characteristics: Gravidity, parity, number of Antenatal Clinic (ANC) contacts, comorbidity, gestational age at delivery (in weeks), previous CD, CD (elective versus emergency), number of fetuses (singleton or multiple), fetal presentation (cephalic, breech, transverse, oblique) or onset of labor (spontaneous, induced or CD before labor began).

Maternal outcomes: Intrapartum (obstructed labor (clinically/labor care guide), prolonged labor), Surgical (Cesarean Hysterectomy, Internal organ injury), Postpartum hemorrhage (estimated blood loss > 500mL or as documented), Other complications (Anemia, wound complications), length of hospital stay (in days) and maternal mortality Neonatal characteristics: Apgar score at 1 minute and 5 minutes (≤3, 4-6, or ≥ 7), Delivery status (live birth or stillbirth), Birthweight in grams

## Potential bias and mitigation measures

This study was potentially subjected to selection bias, as data were obtained from a single referral hospital where complicated pregnancies and high-risk deliveries are more likely to be managed, limiting generalizability to the wider population. In addition, the existence of separate vaginal and caesarean delivery registers could have introduced unequal representation of delivery methods. To minimize this bias, all eligible deliveries recorded during the study were consecutively included from both vaginal and cesarean registers using predefined inclusion criteria.

The study was also vulnerable to information bias due to reliance on retrospective hospital records, which may contain incomplete, inaccurate or inconsistently documented information. This was minimized using an extraction tool which was pre-tested for reliability and validity, cross-checking information across multiple hospital records where possible and excluding records with missing key variables such as maternal age, parity, and mode of delivery.

Confounding bias was another limitation, as factors such as maternal comorbidities, gestational age, previous cesarean section and antenatal care attendance could independently influence obstetric outcomes. To address this, multivariate logistic regression analysis was performed to adjust for potential confounders during statistical analysis.

## Statistical analysis

Data were entered, cleaned and analyzed using STATA MP version 17. Quantitative variables such as maternal age and gestational age were initially analyzed as continuous variables using frequencies and percentages. For analytical purposes, maternal age was categorized into three clinically relevant groups (<20 years, 20 – 34 years, and ≥35 years) based on established obstetric risk classifications commonly used in maternal health research. Parity was similarly grouped into nulliparous, multiparous and grand multiparous categories to facilitate comparison of obstetric outcomes across parity strata.

Descriptive statistics were used to summarize the participants’ demographic, obstetric, and neonatal characteristics. Categorical variables were summarized using frequencies and percentages. Associations between maternal characteristics and obstetric outcomes were initially assessed using Pearson’s chi-square test. Variables with statistical significance in univariate analysis and clinically relevant variables were included in a multivariate logistic regression model to control for confounding factors. Adjusted odds ratios (AORs) with 95% confidence intervals were reported, and a p-value of <0.05 was considered statistically significant.

Subgroup analyses were performed according to maternal age categories and parity groups to examine differences in obstetric and neonatal outcomes across these strata. Interaction between the selected variables, including maternal age and parity, was explored during regression analysis. Missing data were addressed through complete-case analysis. Records of missing information on key study variables, including maternal age, parity and mode of delivery were excluded during data cleaning to minimize information bias and ensure analytical consistency. Sensitivity analyses were not performed because only records with complete and verified data were included in the final analysis.

## Results

### Background characteristics of participants

A total of 1732 delivery records were initially accessed from delivery registers, admission files, and neonatal records. Following data cleaning, verification, and application of study eligibility criteria only 302 were included in the final analysis (fig 1). The mean age of the participants was 28.09 years (SD 6.68). Most of the participants were of optimal reproductive age (68.87%), 134 (44.37%) had secondary education, 39.40% of pregnant women were self employed and 170 (56.29%) were married (Table 1).

**Figure 1:**
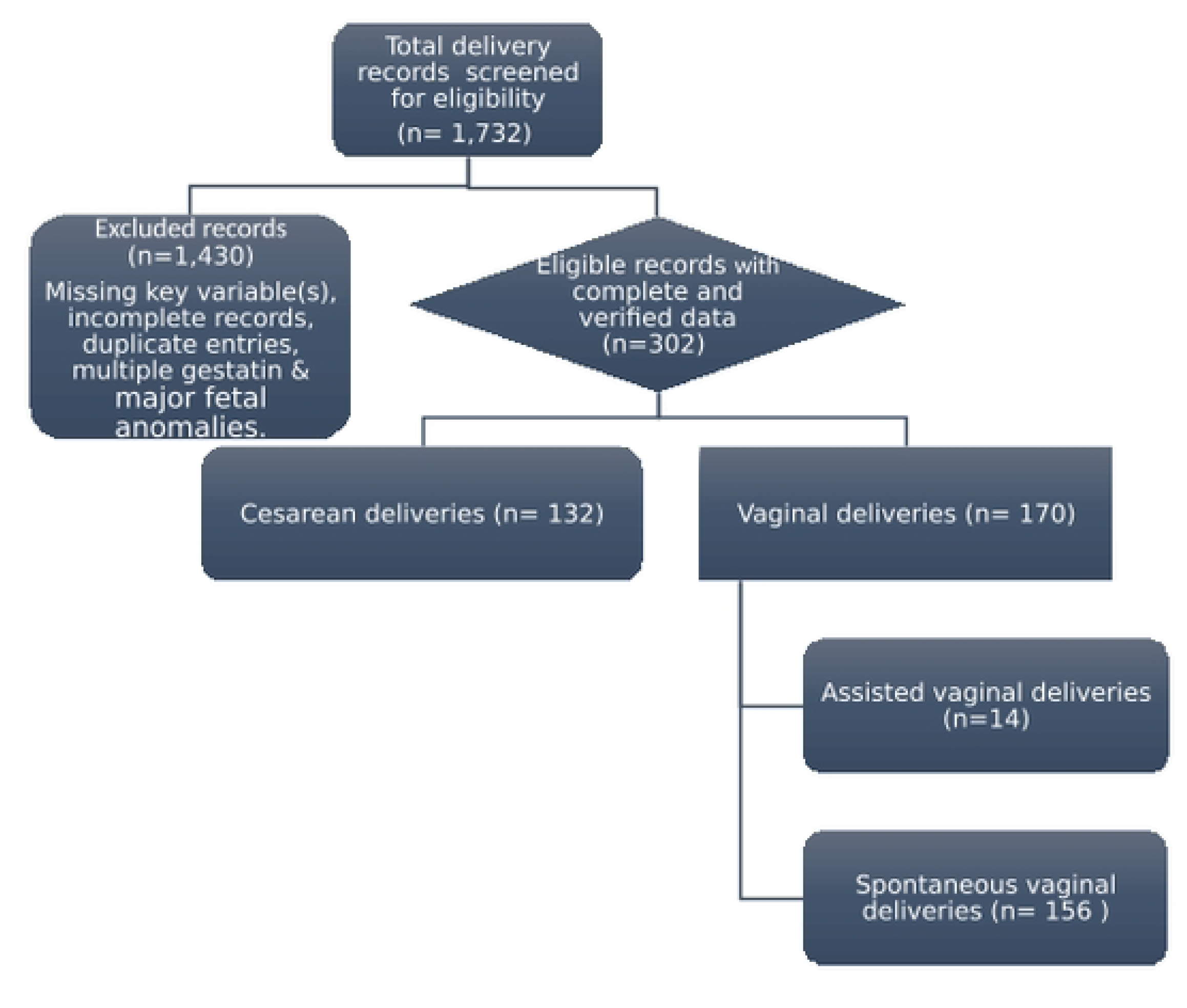
Participants’ flow from screened records to final analytical sample.

**Table 1:**
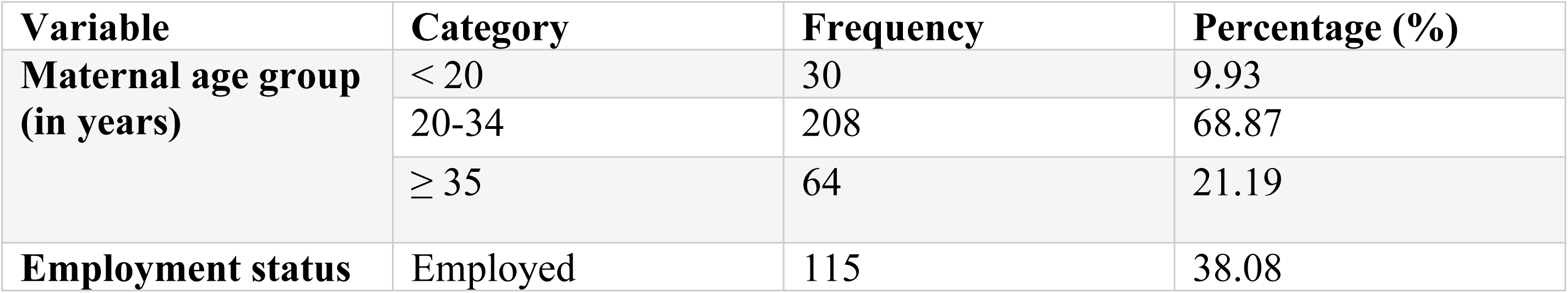
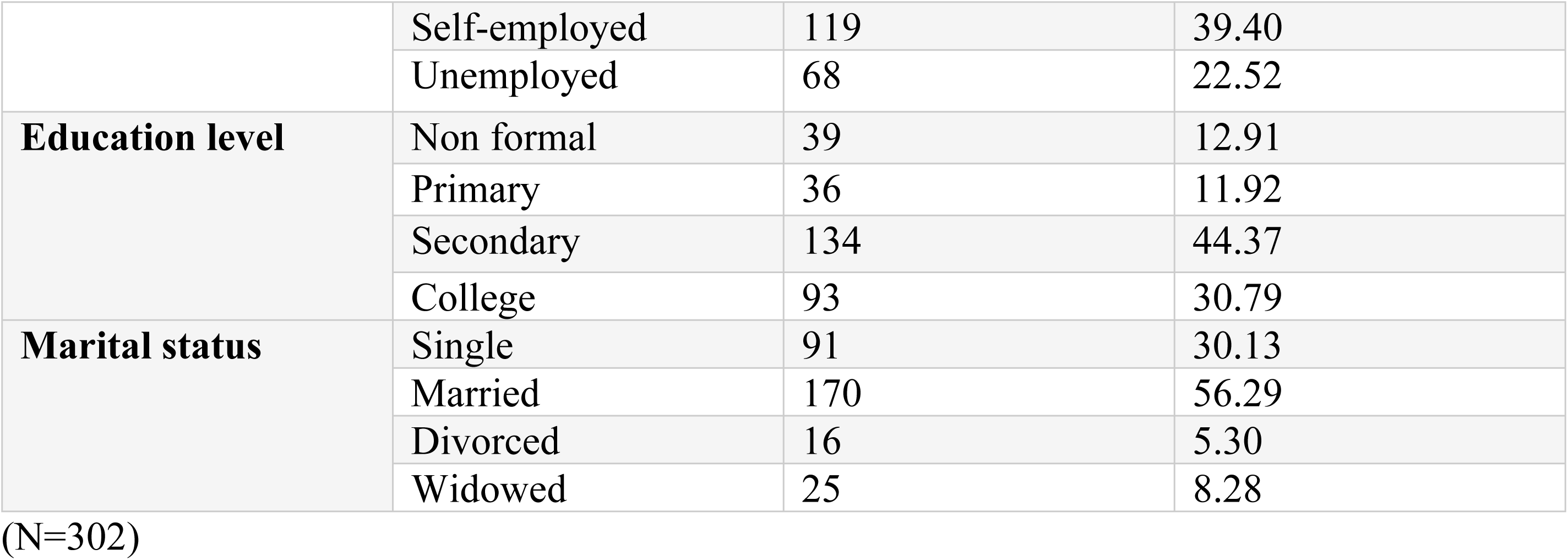
Socio-demographic characteristics of the participants

### Obstetric characteristics of the participants

Most of the participants (59.60%) were multigravidas with more than half classified as multiparous (53.31%) while 96.70% had 1 to 4 living children. 292 (96.69%) had poor antenatal attendance, most had no previous scar (24.17%) while the majority of participants (79.47%) did not have a high-risk pregnancy, nearly half of participants delivered at full term (38.08%) (Table 2).

**Table 2:** Obstetric parameters of participants

| Variables | Category | Frequency | Percentage (%) |
|---|---|---|---|
| **Gravidity** | Primigravida | 96 | 31.79 |
|  | Multigravida | 180 | 59.60 |
|  | Grand multigravida | 26 | 8.61 |
| **Parity** | Nulliparous | 03 | 0.99 |
|  | Primiparous | 125 | 41.39 |
|  | Multiparous | 161 | 53.31 |
|  | Grand multiparous | 13 | 4.30 |
| **Number of living children** | None | 06 | 1.99 |
|  | 1-4 | 286 | 96.70 |
|  | ≥5 | 10 | 3.31 |
| **ANC contacts** | Poor (<8) | 292 | 96.69 |
|  | Recommended (≥8) | 10 | 3.31 |
| **Number of previous scar(s)** | Not applicable (vaginal delivery) | 170 | 56.29 |
|  | None | 73 | 24.17 |
|  | Single | 56 | 18.54 |
|  | Multiple | 03 | 0.99 |
| **High risk pregnancy** | None | 240 | 79.47 |
|  | Antepartum hemorrhage | 15 | 4.97 |
|  | Gestational Diabetes | 07 | 2.32 |
|  | Hypertensive disorders | 26 | 8.61 |
|  | Anemia | 14 | 4.64 |
| **Delivery gestational age (in weeks)** | Preterm | 59 | 19.54 |
|  | Early term | 107 | 35.43 |
|  | Full term | 115 | 38.08 |
|  | Late term | 17 | 5.63 |
|  | Post term | 04 | 1.32 |

(N=302)

### Maternal delivery and outcomes

More than half of the participants (56.29%) had vaginal deliveries including spontaneous vaginal delivery (51.66%) and assisted vaginal delivery (4.64%; vacuum & forceps). The majority of participants (81.79%) had spontaneous onset of labor (Table 3). Most participants had no complications either intrapartum (73.51%) or postpartum (84.11%). However, postpartum hemorrhage was more common among participants of advanced maternal age (15.62%) than among those in other age groups (Table 4).

**Table 3:**
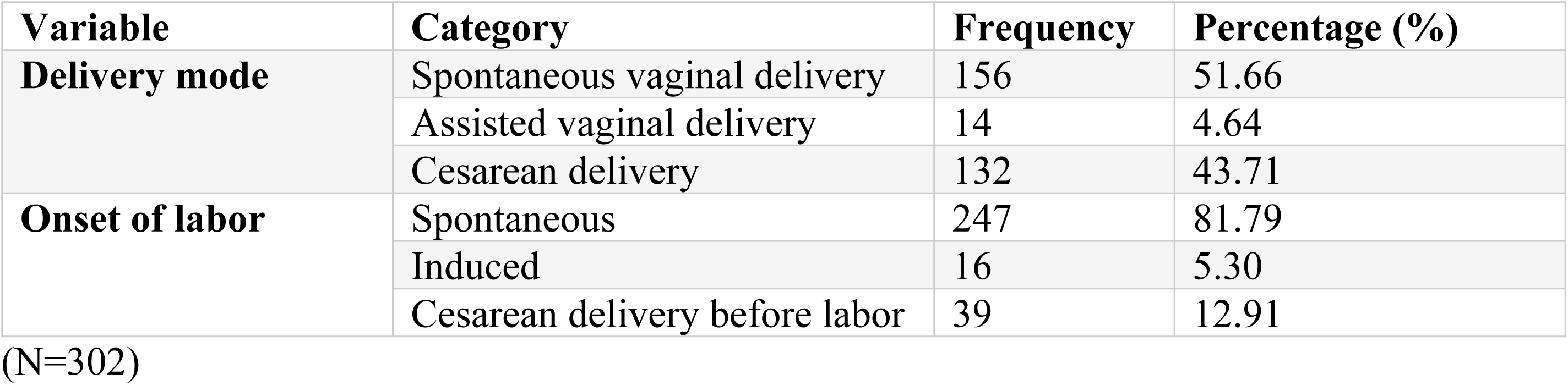
Maternal delivery patterns

**Table 4:** complications among age groups

| **Complications** |  | Maternal age groups (in years) |  |  |
|---|---|---|---|---|
|  |  | <20 | 20-34 | ≥35 |
| **Antepartum** | None | 29 | 198 | 60 |
|  | Antepartum hemorrhage | 1 | 10 | 4 |
| **Intrapartum** | None | 22 | 152 | 48 |
|  | Anesthesia related | 2 | 23 | 08 |
|  | Perineal tear | 05 | 23 | 06 |
|  | Hypertensive disorders | 01 | 08 | 01 |
|  | Eclampsia | 00 | 02 | 01 |
| **Postpartum** | None | 28 | 175 | 51 |
|  | Postpartum hemorrhage | 02 | 24 | 10 |
|  | Hypertensive disorders | 0 | 03 | 0 |
|  | Anemia | 0 | 01 | 01 |
|  | Wound complication | 0 | 05 | 02 |

(N=302)

### Neonatal outcomes

Most of the participants’ newborns had an APGAR score of ≥7 at 1 min (86.42%) and at 5 min (90.73%) with 279(92.38%) live births, while the majority of newborns (81.42%) had a normal birth weight (Table 5).

**Table 5:** Neonatal outcomes

| **Variable** | **Category** | **Frequency** | **Percentage (%)** |
|---|---|---|---|
| **Apgar score at 1 min** | ≤ 3 | 27 | 8.94 |
|  | 4-6 | 14 | 4.64 |
|  | ≥7 | 261 | 86.42 |
| **Apgar score at 5 min** | ≤ 3 | 23 | 7.62 |
|  | 4-6 | 05 | 1.66 |
|  | ≥7 | 274 | 90.73 |
| **Delivery status** | Alive | 279 | 92.38 |
|  | Stillbirth | 23 | 7.68 |
| **Birth weight (in grams)** | ELBW(<1000g) | 04 | 1.32 |
|  | VLBW(<1500g) | 06 | 1.99 |
|  | LBW(<2500g) | 46 | 15.23 |
|  | NBW(≥2500g) | 246 | 81.46 |

(N=302)

**Table 6:**
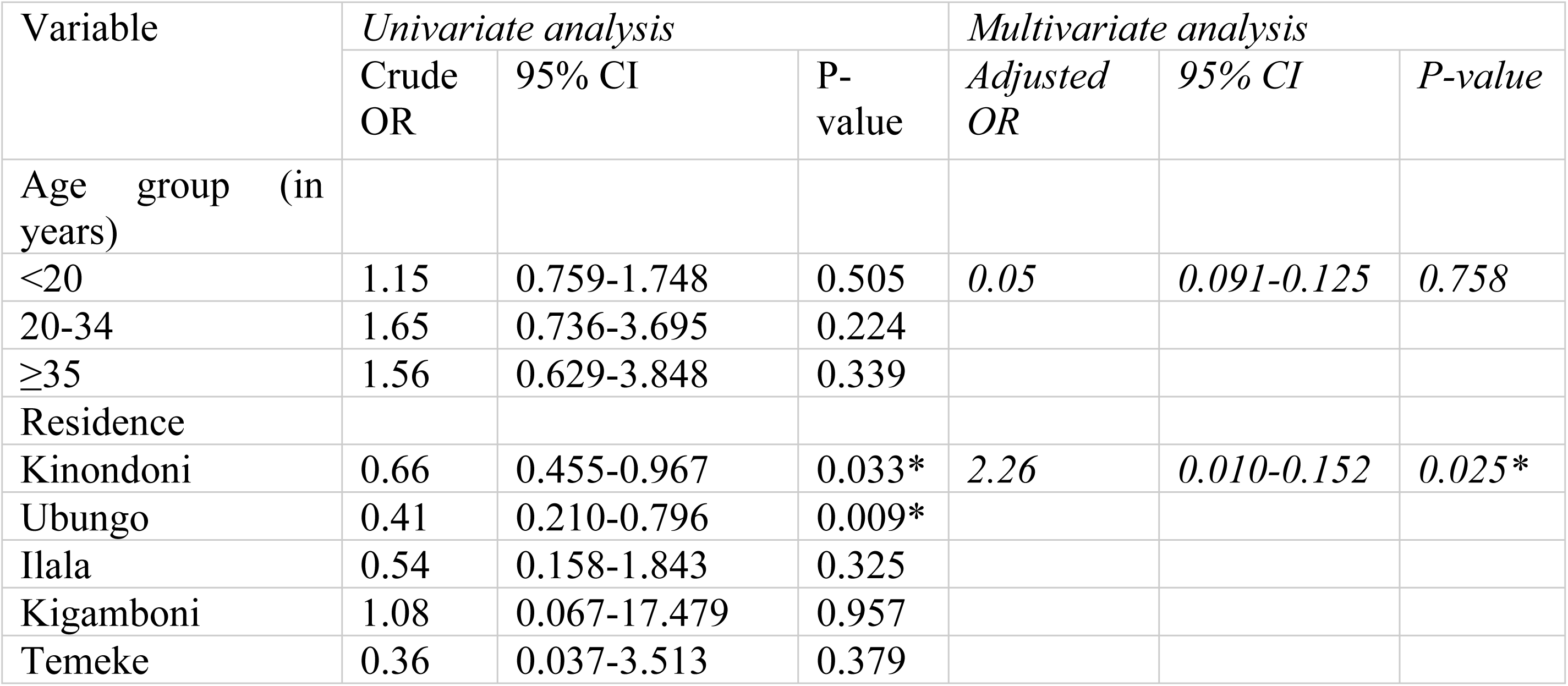
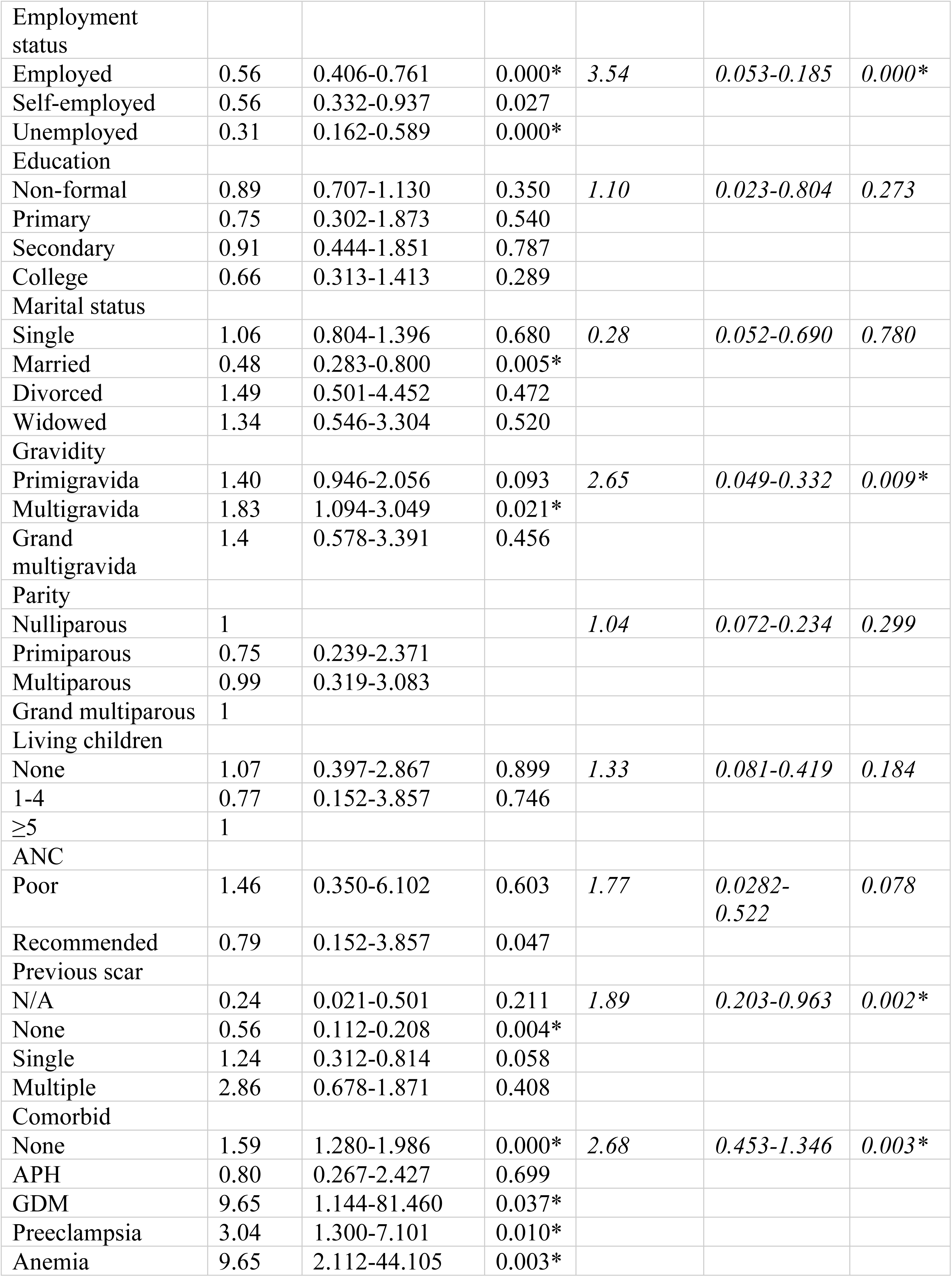
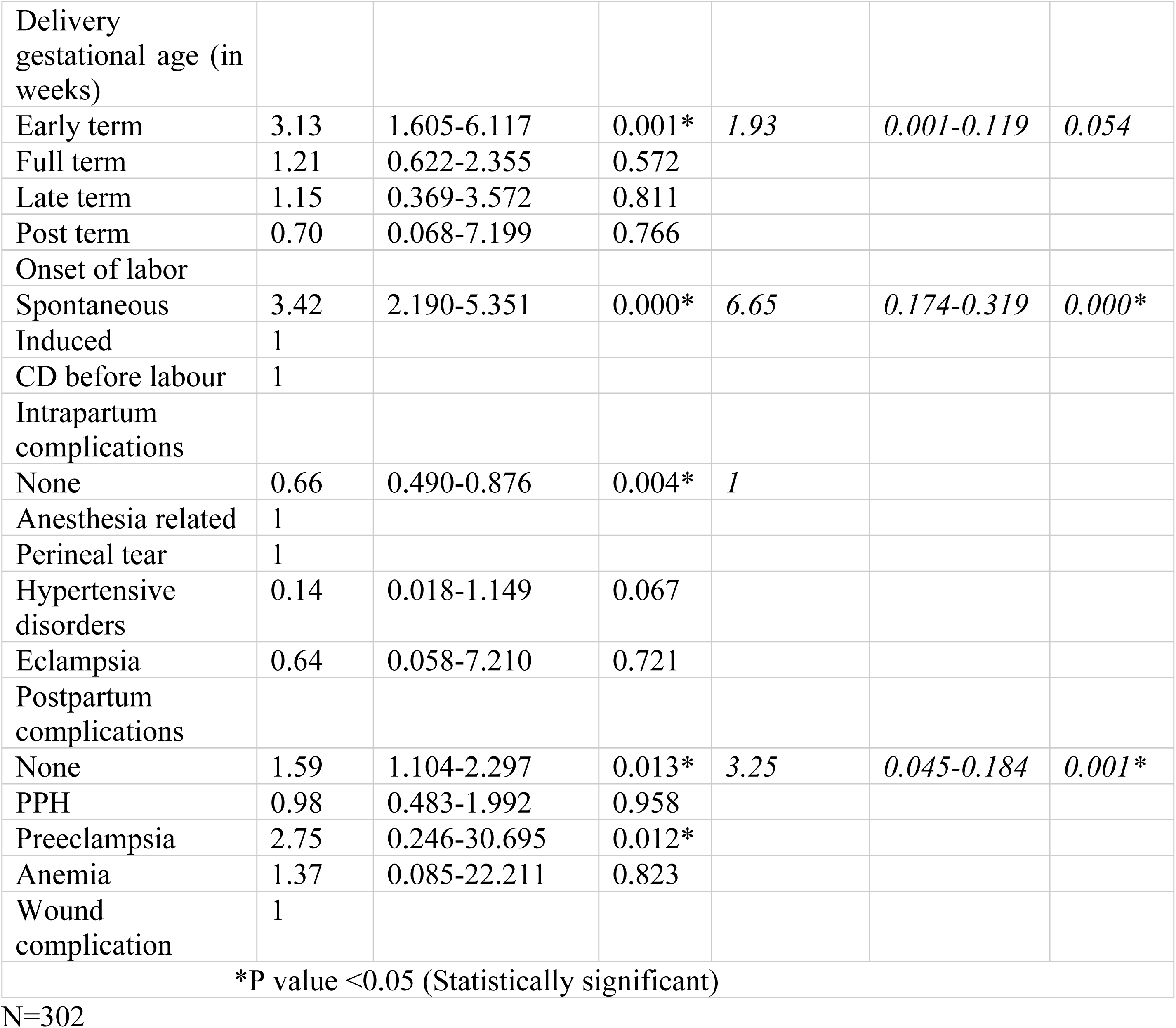
Factors associated with Cesarean delivery

| Variable | *Univariate analysis* |  |  | *Multivariate analysis* |  |  |
|---|---|---|---|---|---|---|
|  | Crude OR | 95% CI | P-value | *Adjusted OR* | *95% CI* | *P-value* |
| Age group (in years) |  |  |  |  |  |  |
| <20 | 1.15 | 0.759-1.748 | 0.505 | *0.05* | *0.091-0.125* | *0.758* |
| 20-34 | 1.65 | 0.736-3.695 | 0.224 |  |  |  |
| ≥35 | 1.56 | 0.629-3.848 | 0.339 |  |  |  |
| Residence |  |  |  |  |  |  |
| Kinondoni | 0.66 | 0.455-0.967 | 0.033* | *2.26* | *0.010-0.152* | *0.025\** |
| Ubungo | 0.41 | 0.210-0.796 | 0.009* |  |  |  |
| Ilala | 0.54 | 0.158-1.843 | 0.325 |  |  |  |
| Kigamboni | 1.08 | 0.067-17.479 | 0.957 |  |  |  |
| Temeke | 0.36 | 0.037-3.513 | 0.379 |  |  |  |
| Employment status |  |  |  |  |  |  |
| Employed | 0.56 | 0.406-0.761 | 0.000* | *3.54* | *0.053-0.185* | *0.000\** |
| Self-employed | 0.56 | 0.332-0.937 | 0.027 |  |  |  |
| Unemployed | 0.31 | 0.162-0.589 | 0.000* |  |  |  |
| Education |  |  |  |  |  |  |
| Non-formal | 0.89 | 0.707-1.130 | 0.350 | *1.10* | *0.023-0.804* | *0.273* |
| Primary | 0.75 | 0.302-1.873 | 0.540 |  |  |  |
| Secondary | 0.91 | 0.444-1.851 | 0.787 |  |  |  |
| College | 0.66 | 0.313-1.413 | 0.289 |  |  |  |
| Marital status |  |  |  |  |  |  |
| Single | 1.06 | 0.804-1.396 | 0.680 | *0.28* | *0.052-0.690* | *0.780* |
| Married | 0.48 | 0.283-0.800 | 0.005* |  |  |  |
| Divorced | 1.49 | 0.501-4.452 | 0.472 |  |  |  |
| Widowed | 1.34 | 0.546-3.304 | 0.520 |  |  |  |
| Gravidity |  |  |  |  |  |  |
| Primigravida | 1.40 | 0.946-2.056 | 0.093 | *2.65* | *0.049-0.332* | *0.009\** |
| Multigravida | 1.83 | 1.094-3.049 | 0.021* |  |  |  |
| Grand multigravida | 1.4 | 0.578-3.391 | 0.456 |  |  |  |
| Parity |  |  |  |  |  |  |
| Nulliparous | 1 |  |  | *1.04* | *0.072-0.234* | *0.299* |
| Primiparous | 0.75 | 0.239-2.371 |  |  |  |  |
| Multiparous | 0.99 | 0.319-3.083 |  |  |  |  |
| Grand multiparous | 1 |  |  |  |  |  |
| Living children |  |  |  |  |  |  |
| None | 1.07 | 0.397-2.867 | 0.899 | *1.33* | *0.081-0.419* | *0.184* |
| 1-4 | 0.77 | 0.152-3.857 | 0.746 |  |  |  |
| ≥5 | 1 |  |  |  |  |  |
| ANC |  |  |  |  |  |  |
| Poor | 1.46 | 0.350-6.102 | 0.603 | *1.77* | *0.0282-0.522* | *0.078* |
| Recommended | 0.79 | 0.152-3.857 | 0.047 |  |  |  |
| Previous scar |  |  |  |  |  |  |
| N/A | 0.24 | 0.021-0.501 | 0.211 | *1.89* | *0.203-0.963* | *0.002\** |
| None | 0.56 | 0.112-0.208 | 0.004* |  |  |  |
| Single | 1.24 | 0.312-0.814 | 0.058 |  |  |  |
| Multiple | 2.86 | 0.678-1.871 | 0.408 |  |  |  |
| Comorbid |  |  |  |  |  |  |
| None | 1.59 | 1.280-1.986 | 0.000* | *2.68* | *0.453-1.346* | *0.003\** |
| APH | 0.80 | 0.267-2.427 | 0.699 |  |  |  |
| GDM | 9.65 | 1.144-81.460 | 0.037* |  |  |  |
| Preeclampsia | 3.04 | 1.300-7.101 | 0.010* |  |  |  |
| Anemia | 9.65 | 2.112-44.105 | 0.003* |  |  |  |
| Delivery gestational age (in weeks) |  |  |  |  |  |  |
| Early term | 3.13 | 1.605-6.117 | 0.001* | *1.93* | *0.001-0.119* | *0.054* |
| Full term | 1.21 | 0.622-2.355 | 0.572 |  |  |  |
| Late term | 1.15 | 0.369-3.572 | 0.811 |  |  |  |
| Post term | 0.70 | 0.068-7.199 | 0.766 |  |  |  |
| Onset of labor |  |  |  |  |  |  |
| Spontaneous | 3.42 | 2.190-5.351 | 0.000* | *6.65* | *0.174-0.319* | *0.000\** |
| Induced | 1 |  |  |  |  |  |
| CD before labour | 1 |  |  |  |  |  |
| Intrapartum complications |  |  |  |  |  |  |
| None | 0.66 | 0.490-0.876 | 0.004* | *1* |  |  |
| Anesthesia related | 1 |  |  |  |  |  |
| Perineal tear | 1 |  |  |  |  |  |
| Hypertensive disorders | 0.14 | 0.018-1.149 | 0.067 |  |  |  |
| Eclampsia | 0.64 | 0.058-7.210 | 0.721 |  |  |  |
| Postpartum complications |  |  |  |  |  |  |
| None | 1.59 | 1.104-2.297 | 0.013* | *3.25* | *0.045-0.184* | *0.001\** |
| PPH | 0.98 | 0.483-1.992 | 0.958 |  |  |  |
| Preeclampsia | 2.75 | 0.246-30.695 | 0.012* |  |  |  |
| Anemia | 1.37 | 0.085-22.211 | 0.823 |  |  |  |
| Wound complication | 1 |  |  |  |  |  |
| *P value <0.05 (Statistically significant) |  |  |  |  |  |  |

N=302

### Factors associated with Cesarean delivery

Employment status, marital status, gravidity, previous scar, comorbidities, delivery gestational age, onset of labor, intrapartum complications and postpartum complications were the determinants of caesarian delivery (P< 0.05).

Those who were employed had three times the likelihood of cesarean delivery (AOR=3.54, 95% CI 0.053-0.185, P=0.000), being married was associated with half the likelihood of cesarean delivery compared to others (single, divorced, widowed) (OR=0.48, 95% CI 0.233-0.800, P=0.005). Primigravida had more than twice the likelihood of cesarean delivery compared to multigravida (AOR=2.65, 95% CI 0.049-0.332, P=0.009), previous scar was associated with nearly twice the likelihood of cesarean delivery compared to those with none (AOR=1.89, 95% CI 0.203-0.963, P=0.002), comorbidities contributed to three times the chance of cesarean delivery compared to those who had none (AOR=2.68, 95% CI 0.453-1.346, P=0.000).

Early term deliveries had a threefold likelihood of a cesarean delivery compared to other gestational ages (OR=3.13, 95% CI 1.605-6.117, P=0.001), spontaneous onset of labor increased cesarean delivery to about seven times compared to those who had before the onset of labor (AOR=6.65, 95% CI 0.174-0.319, P=0.000), having neither intrapartum (AOR=1, P=0.004) nor postpartum complication (AOR=3.25, 95% CI 0.045-0.184, P=0.001) was linked to cesarean delivery.

## Discussion

This study examined caesarean delivery patterns and selected maternal and neonatal outcomes among women delivering at Mwananyamala Regional Referral Hospital (MRRH) in Dar es Salaam, Tanzania. The findings demonstrated a high caesarean delivery rate, a relatively high proportion of mothers at the extremes of reproductive age, and generally favorable neonatal outcomes despite persistent risks of postpartum hemorrhage, low birth weight, and stillbirth. Several maternal sociodemographic and obstetric factors were independently associated with caesarean delivery.

The distribution of maternal age showed that 9.93% of mothers were adolescents (<20 years while, 21.19% were of advanced maternal age (≥35 years). These findings are consistent with reports from Sub-Saharan Africa where adolescent pregnancies remain common with births exceeding 90 per 1,000 population, and WHO data indicate that the proportion of pregnancies among older women is on the rise, (≈12.3% of pregnancies in developing countries occur in women ≥35) due to delayed childbearing and changing fertility patterns. Similar studies from Tanzania also reported 16% of deliveries to women of advanced age[6], although variations may occur because of differences in urbanization, access to reproductive health services, and referral patterns. The relatively high proportion of older mothers observed in this study may reflect the urban population served by MRRH and its role as a referral center for high-risk pregnancies. Since maternal age extremes are well known to carry higher obstetric risk[14], these findings emphasize the importance of targeted antenatal surveillance and individualized obstetric care for women at extremes of reproductive age.

Most women were parous (53.3% multiparous), and only 4.3% were grand-multiparous. This prevalence of grand multiparity was lower than that reported in Northern Tanzania (≈9.4%)[15]. This difference might reflect more urban fertility trends, a coastal rather than lake zone lifestyle or sampling. Nonetheless, high-parity women are recognized to have elevated risks (e.g. for hemorrhage and perinatal loss)[8]. These findings imply that the obstetric population attending Mwananyamala Regional Referral Hospital (MRRH) includes high-risk groups (young and old mothers, as well as multiparous women), highlighting the need for age and parity-tailored antenatal counselling and monitoring.

The caesarean delivery rate observed in this study (43.71%) was substantially higher than the World Health Organization recommended range of 10-15% and exceeded the Tanzanian national average(≈13%)[16] and pooled estimates from East Africa (≈24%)[17]. Similarly elevated caesarean delivery rates have been reported in tertiary and referral hospitals managing complicated pregnancies and obstetric emergencies. The high rate observed at MRRH may therefore reflect referral bias, increasing obstetric risk profiles, and expanding access to surgical obstetric services in urban settings. Although caesarean delivery can be lifesaving when medically indicated, persistently high rates raise concerns regarding unnecessary procedures, increased healthcare costs, intraoperative and postoperative complications. These findings support the need for continuous clinical audit of caesarean delivery indications and implementation of standardized classification systems to promote appropriate use of operative delivery.

This study further showed that primigravity, previous uterine scar, maternal comorbidities, early term pregnancies and employment status were independently associated with caesarean delivery. Similar associations have been reported in studies from Tanzania[16]. Primigravid women are more likely to experience prolonged labor and cephalopelvic disproportion, which may increase the likelihood of operative delivery. Likewise, women with previous uterine scars are commonly managed more cautiously because of the risk of uterine rupture during labor. Maternal comorbidities such as hypertensive disorders and gestational diabetes may also necessitate surgical intervention to reduce maternal and fetal complications. Early-term gestation was associated with higher adjusted odds of caesarean delivery. This may reflect planned or clinically indicated delivery before 39 completed weeks among women with obstetric risk, maternal comorbidities, or fetal concern(..). The association between employment status and caesarean delivery may reflect socioeconomic disparities, differences in healthcare-seeking behavior, or increased preference for higher level facility-based obstetric interventions among employed women. These findings highlight the multifactorial nature of caesarean delivery and the importance of comprehensive antenatal risk assessment.

Severe maternal complications were relatively infrequent across age groups in this study. About 73.5% of women had no intrapartum complications and 84.1% had no postpartum complications. Notably, the rate of postpartum hemorrhage (PPH) appeared higher among women of advanced maternal age (AMA). This study is consistent with previous literature linking advanced maternal age and higher parity to PPH[8]. In contrast, hypertensive disorders such as preeclampsia and eclampsia were uncommon across all age groups. This is lower than some reports of excess preeclampsia in AMA[6] suggesting effective prenatal care by MRRH, a different sampling area or a small sample size.

Newborn outcomes were generally favorable, 92.4% of infants were live births, and most neonates achieving APGAR scores ≥7 at both one and five minutes. However, the prevalence of low birth weight (15.23%) and stillbirth (7.68%) remains clinically significant. Similar findings have been reported from some Tanzanian tertiary centers; a five-year Dar es Salaam audit found 7% stillbirths and 21% low-birth weight[18], and a northern Tanzanian registry reported increased odds of low birthweight and stillbirth with maternal age[6]. These adverse outcomes may be linked to maternal comorbidities, preterm delivery, and obstetric complications, particularly among women at the extremes of reproductive age. The findings suggest that although obstetric and neonatal care at MRRH is generally effective, continued strengthening of fetal monitoring, neonatal resuscitation, and high-risk pregnancy management is necessary to further reduce preventable neonatal morbidity and mortality.

## Study limitations

First the retrospective design relied on secondary hospital records, making the study susceptible to incomplete documentation and information bias. Second the exclusion of records with missing data may have introduced selection bias and reduced the representativeness of the final sample. Third, because the study was conducted at a single urban referral hospital, the findings may not be generalizable to rural settings or lower-level health facilities. Finally, causal relationship could not be established because of the cross-sectional retrospective nature of the study. Despite these limitations, the study provides important baseline evidence on caesarean delivery patterns and selected maternal and neonatal outcomes in an urban Tanzanian referral setting.

## Conclusion

Most mothers were aged 20-34 years, although adolescent and advanced maternal age pregnancies remained common. The caesarean section rate was high and was independently associated with primigravity, previous uterine scar, maternal comorbidities, and early term gestation. While maternal and neonatal outcomes were generally favorable, targeted efforts to improve hemorrhage prevention and ensure appropriate indications for caesarean delivery are needed.

## Data Availability

All data are available

## References

1. Maternal mortality rates and statistics. UNICEF DATA n.d. https://data.unicef.org/topic/maternal-health/maternal-mortality/ (accessed July 15, 2025).

2. Neonatal mortality. UNICEF DATA n.d. https://data.unicef.org/topic/child-survival/neonatal-mortality/ (accessed July 15, 2025).

[3] WHO. Adolescent pregnancy. World Health Organ 2024. https://www.who.int/news-room/fact-sheets/detail/adolescent-pregnancy (accessed July 15, 2025).

4. The Social and Educational Consequences of Adolescent Childbearing - World Bank Gender Data Portal. World Bank Gend Data Portal n.d. http://genderdata.worldbank.org/en/data-stories/adolescent-fertility (accessed July 15, 2025).

[5] Laopaiboon M, Lumbiganon P, Intarut N, Mori R, Ganchimeg T, Vogel J, et al. Advanced maternal age and pregnancy outcomes: a multicountry assessment. BJOG Int J Obstet Gynaecol 2014;121:49–56. 10.1111/1471-0528.12659.

[6] Mnabwiru LR, Cho YS, Mahande MJ, Mazugun N, Mchome BL, Park EY. Impact of advanced maternal age on perinatal outcomes in Tanzania: Insights from Kilimanjaro Christian Medical Center Birth Registry. Heliyon 2024;10:e24608. 10.1016/j.heliyon.2024.e24608.

[7] Vandekerckhove M, Guignard M, Civadier M-S, Benachi A, Bouyer J. Impact of maternal age on obstetric and neonatal morbidity: a retrospective cohort study. BMC Pregnancy Childbirth 2021;21:732. 10.1186/s12884-021-04177-7.

[8] Ogbe AE, Ogbe BP, Ekwempu C. Obstetric Outcome in Grandmultiparous Women in Jos University Teaching Hospital. Jos J Med 2012;6:39–43.

[9] Mutihir JT. Obstetric outcome of the grandmultipara in Jos, Nigeria. J Med Trop 2005;7:14–20. 10.4314/jmt.v7i1.35178.

[10] Eze JN, Okaro JM, Okafor MH. Outcome of pregnancy in the Grandmultipara in Enugu, Nigeria. Trop J Obstet Gynaecol 2006;23:8–11. 10.4314/tjog.v23i1.14555.

[11] Cresswell J. Trends in Maternal Mortality 2000 to 2020: Estimates by WHO, UNICEF, UNFPA, World Bank Group and UNDESA/Population Division. 1st ed. Geneva: World Health Organization; 2023.

[12] Leslie K. Survey Sampling. New York: John Wiley and Sons. 1965:78–94.

[13] Kibe PM, Mbuthia GW, Shikuku DN, Akoth C, Oguta JO, Ng L, et al. Prevalence and factors associated with caesarean section in Rwanda : a trend analysis of Rwanda demographic and health survey 2000 to 2019 – 20. BMC Pregnancy Childbirth 2022:1–13. 10.1186/s12884-022-04679-y.

[14] Amoadu M, Hagan D, Ansah EW. Adverse obstetric and neonatal outcomes of adolescent pregnancies in Africa: a scoping review. BMC Pregnancy Childbirth 2022;22:598. 10.1186/s12884-022-04821-w.

[15] Muniro Z, Tarimo CS, Mahande MJ, Maro E, Mchome B. Grand multiparity as a predictor of adverse pregnancy outcome among women who delivered at a tertiary hospital in Northern Tanzania. BMC Pregnancy Childbirth 2019;19:222. 10.1186/s12884-019-2377-5.

[16] Nahayo B, Olorunfemi G, Ndayishimye S, Nsanzabera C. Prevalence and factors associated with caesarean section among Tanzanian women of reproductive age: evidence from the 2022 Tanzania demographic and health survey data. BMC Public Health 2025;25:794. 10.1186/s12889-025-21967-2.

[17] Habteyes AT, Mekuria MD, Negeri HA, Kassa RT, Deribe LK, Sendo EG. Prevalence and associated factors of caesarean section among mothers who gave birth across Eastern Africa countries: Systematic review and meta-analysis study. Heliyon 2024;10. 10.1016/j.heliyon.2024.e32511.

[18] Kamala BA, Mgaya AH, Ngarina MM, Kidanto HL. Predictors of low birth weight and 24- hour perinatal outcomes at Muhimbili National Hospital in Dar es Salaam, Tanzania: a five- year retrospective analysis of obstetric records. Pan Afr Med J 2018;29. 10.11604/pamj.2018.29.220.15247.

